# Impairment in acquisition of conditioned fear in schizophrenia: a pooled analysis of four studies

**DOI:** 10.1101/2021.05.26.21257857

**Authors:** Lauri Tuominen, Liana Romaniuk, Mohammed R. Milad, Donald C. Goff, Jeremy Hall, Daphne J. Holt

## Abstract

**Background:** Individuals with schizophrenia show impairments in associative learning. One well-studied, quantifiable form of associative learning is Pavlovian fear conditioning. However, to date, studies of fear conditioning in schizophrenia have been inconclusive, possibly because they lacked sufficient power.

**Methods:** To address this issue, data were pooled from 4 independent fear conditioning studies that included a total of 77 individuals with schizophrenia and 74 control subjects. Skin conductance responses (SCRs) during fear conditioning to stimuli that were paired (the CS+) and not paired (CS-) with an aversive, unconditioned stimulus were measured, and the success of acquisition of differential conditioning (the magnitude of CS+ vs. CS-SCRs) and responses to CS+ and CS-separately were assessed.

**Results:** Acquisition of differential conditioned fear responses was significantly lower in individuals with schizophrenia than in healthy controls (Cohen’s d = 0.53). This effect was primarily related to a significantly higher response to the CS-stimulus in the schizophrenia compared to the control group. The magnitude of this response to the CS- in the schizophrenia group was correlated with the severity of delusional ideation. Other symptoms or antipsychotic dose were not associated with fear conditioning measures.

**Conclusions:** Individuals with schizophrenia who endorse delusional beliefs are over-responsive to neutral stimuli during fear conditioning. This finding is consistent with prior models of aberrant learning in psychosis.

## Introduction

An impairment in associative learning may represent a critical step on the path linking genetic and environmental risk factors for schizophrenia to the formation both positive and negative symptoms^1^. For instance, delusional symptoms of schizophrenia have been suggested to arise from incorrected inferences from evidence^2^. The influential aberrant salience attribution theory of psychosis ^3^ posits that individuals with schizophrenia assign meaning to neutral or irrelevant events and internal representations. According to this theory, false beliefs and ultimately delusions arise from attempts to explain why previously neutral or irrelevant events suddenly command attention and hold meaning. On the other hand, schizophrenia has also been associated with diminished responses to and learning from salient stimuli, such as rewards^4^. This impairment in turn could explain persistent deficits in motivation and social functioning in schizophrenia. Thus, both inappropriate and deficient assignments of salience have been associated with schizophrenia and may potentially underlie positive and negative symptoms, respectively. However, there remains limited direct empirical support for these models.

Pavlovian fear conditioning is one simple experimental paradigm that can be used to quantitatively measure associative learning^5^ and test such models directly in humans. In a typical fear conditioning experiment, two distinct stimuli are presented: 1) a neutral stimulus (often a visual image in studies conducted in humans) that is repeatedly paired with an aversive unconditioned stimulus (US), such as an electrical shock, unpleasant picture or loud noise, the CS+, and 2) another neutral stimulus that is never paired with the US, the CS-. Over the course of the experiment, subjects learn to associate the paring of US with the CS+. As a result, subjects show similar responses to the CS+ that were initially elicited by the US, for example a galvanic skin conductance response. Thus, the main outcome measure, reflecting the success of acquisition of conditioned fear responses and the ability to form associations, is the difference between the responses to the CS+ and the CS- stimuli. A hypothesis derived from the aberrant salience attribution theory of psychosis proposes that, due to increases in spontaneous dopamine release, individuals with schizophrenia exhibit increased responses to the neutral stimulus (i.e, the CS-) during fear conditioning ^3,6,7^. Conversely, the proposed decrease in relevant cue-induced adaptive hypofunction of the dopamine systems would result in a decreased response to the CS+ stimulus in schizophrenia ^7^. Together, the expected pattern, increased responses to neutral stimuli and decreased responses to the conditioned stimuli, would result in impaired acquisition of differential conditioned fear responses in schizophrenia.

Although schizophrenia has been studied extensively using Pavlovian conditioning paradigms, no consistent picture has yet emerged ^6,8–16^. Some studies have found evidence for diminished differences between responses to CS+ and CS-stimuli in schizophrenia and a higher response to the CS-^6,8,10^, whereas other studies have not found statistically significant differences^9,12^. These discrepant findings may be in part due to the small sample sizes in these previously published studies. Also, clinical features associated with the putative deficits in acquisition of conditioned fear have not been identified.

Thus, here we aimed to address some of these unresolved questions by pooling data from four previously published studies on fear conditioning in schizophrenia. We limited this pooling to research published in the past 20 years in order to maximize the similarity in methods and definitions of schizophrenia, as well as the availability of the data, across the studies. All four studies^8–10,12^ that were included in the analyses had collected skin conductance responses to CS+ and CS-stimuli during a Pavlovian fear conditioning paradigm in individuals diagnosed with schizophrenia and healthy control subjects and measured symptom severity using standard instruments in the schizophrenia group. In this pooled sample of 77 individuals with schizophrenia and 74 healthy control subjects, we tested whether impaired acquisition of differential fear conditioning was evident in the schizophrenia, compared to the healthy control group. We then tested the following specific predictions: Compared to the controls, the participants of the schizophrenia group would show: 1) higher responses to the CS-stimulus, which would be associated with more severe delusional ideation, and 2) lower responses to the CS+ stimulus, which would associated with more severe negative symptoms.

## Methods

### The sample

One study^6^ with relevant data was not included because the original data were no longer available. Data of four independent studies were included in the analyses, collected from a total of 77 individuals with schizophrenia and 74 healthy control subjects (**Table 1**). Following quality control procedures, one individual with schizophrenia from Holt et al. 2009 ^10^ was excluded from all analyses due to that subject’s outlier status (see below); therefore 76 subjects were included in the final combined schizophrenia group. All four studies defined schizophrenia using DSM-IV criteria which were assessed using a standardized, structured clinical interview ^17^. Symptom severity was measured using the Positive and Negative Syndrome Scale (PANSS)^18^. Antipsychotic medication dose was reported as chlorpromazine equivalents. In all studies, fear conditioning responses were measured using skin conductance. Each individual study had obtained ethics committee approval separately either from the Partners Healthcare Institutional Review Board or Lothian National Health Service Research Ethics Committee.

**Table 1.** Demographic characteristics of the participants of each study.

|  | Holt et al., 2009 |  | Romaniuk et al., 2010 |  | Holt et al., 2012 |  | Tuominen et al., 2021 |  | Combined |  |
| --- | --- | --- | --- | --- | --- | --- | --- | --- | --- | --- |
|  | CTR | SCZ | CTR | SCZ | CTR | SCZ | CTR | SCZ | CTR | SCZ |
| **N** | 15 | 15 | 19 | 20 | 15 | 18 | 25 | 23 | 74 | 76 |
| **Age (years)** | 41.20<br>(14.75) | 43.93<br>(11.70) | 35.42<br>(8.28) | 36.35<br>(9.40) | 34.80<br>(10.16) | 34.67<br>(9.50) | 29.00<br>(6.16) | 29.96<br>(6.81) | 34.30<br>(10.52) | 35.51<br>(10.29) |
| **Sex (number/% of males)** | 10 / 66.7 | 10 / 66.7 | 13 / 68.4 | 14 / 70.0 | 15 / 100.0 | 18 / 100.0 | 16 / 64.0 | 17 / 73.9 | 54 / 73.0 | 59 / 77.6 |
| **PANSS Positive Subscale Score** | - | 15.00<br>(4.49) | - | 10.05<br>(2.63) | - | 13.28<br>(5.18) | - | 15.91<br>(5.51) | - | 13.57<br>(5.09) |
| **PANSS Negative Subscale Score** | - | 13.60<br>(3.91) | - | 14.15<br>(3.79) | - | 13.22<br>(5.49) | - | 17.30<br>(5.30) | - | 14.78<br>(4.95) |
| **PANSS Total Score** | - | 58.87<br>(11.22) | - | 48.50<br>(9.34) | - | 52.17<br>(12.81) | - | 64.70<br>(14.97) | - | 56.32<br>(13.85) |
| **CPZ equivalents** | - | 357.70<br>(282.11) | - | 302.40<br>(180.03) | - | 249.22<br>(316.58) | - | 384.47<br>(284.64) | - | 325.56<br>(269.00) |

Table shows mean (SD) values except for sex which shows the number/percentage of males.

The four studies differed in some aspects of their design and methods, including whether the data was collected inside an MRI scanner or not, whether the US was a mild electrical shock or an unpleasant picture, the reinforcement rate, number of trials, duration of stimulus presentation, and duration of the overall experiment. These specific methodological characteristics of the studies are described in Table 2.

**Table 2.**
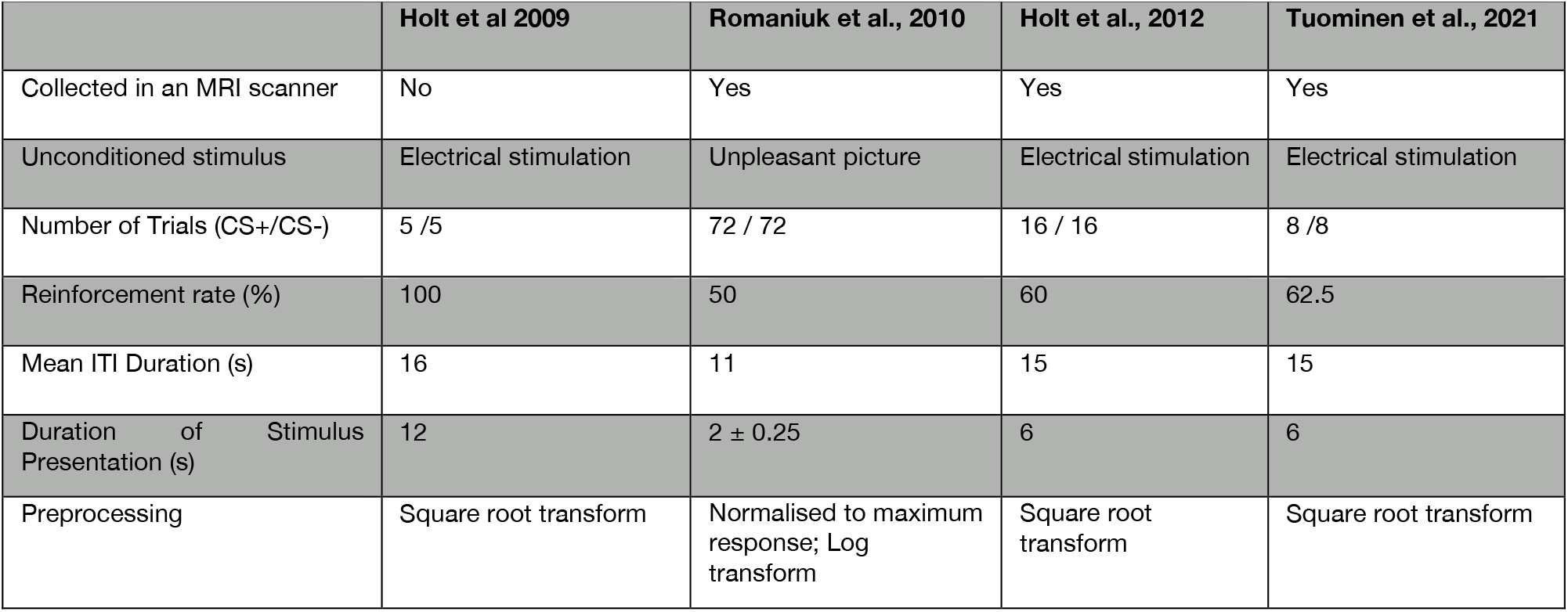
The experimental design and parameters of each study.

### Data quality control

In one participant of the schizophrenia group of the Holt et al., 2009 study, the CS+ vs. CS-contrast was greater than 4 times the interquartile interval from the mean. This subject was therefore excluded from all subsequent statistical analyses.

### Clinical measures

In three out of four studies, delusional symptoms were measured using Peter’s Delusion Inventory (PDI) ^19,20^, which is a self-report questionnaire measuring delusional ideation validated for use in both non-clinical and schizophrenia samples ^21,22^. Two studies used a 40-item version of the PDI^19^ and one study use the 21-item PDI^20^. These data were normalized across studies by dividing the total score by the total number of items. In addition, positive, negative, and general symptoms of schizophrenia, as well as the total burden of symptoms, were assessed in all studies using the Positive and Negative Syndrome Scale (PANSS)^18^. Also, information about treatment with antipsychotic medication (current daily dose) was obtained.

### Statistical tests

#### Primary analyses

Our main hypothesis was that the difference in skin conductance responses between the CS+ and CS-was significantly different between the schizophrenia and control groups (i.e., lower in the schizophrenia group). This hypothesis was tested using a linear mixed effects model: CS+ vs CS-contrast ∼ schizophrenia diagnosis + sex + age. Study (i.e., of the four original cohorts) was entered as a random factor into the model. Separate linear mixed effects models were generated to test whether the two groups differed in response to the CS+ or CS-stimuli separately, while controlling for age and sex, and again including study as a random factor.

In a subset of the schizophrenia group (n=56) who had completed the PDI, we tested if the response to the CS- was correlated with the total PDI score using bi-square robust linear regression, with sex, age and study as covariates. Similarly, we tested in the full schizophrenia sample (n=76) whether the response to the CS+ was correlated with the PANSS negative symptom subscale score, using bi-square robust linear regression, with sex, age and study as covariates.

#### Secondary, exploratory analyses

We explored whether the severity of positive, negative, general, or total symptoms, or current antipsychotic medication dose, were associated with any of the fear conditioning measures in the schizophrenia group, while controlling for age, sex and study.

#### Confirmatory meta-analysis

We also conducted a meta-analysis in order to a) confirm the results of the linear mixed effects model, b) calculate a Cohen’s d value for the combined sample, and c) to obtain an estimate of the heterogeneity of the studies.

We chose to use robust regression since this method provides more accurate estimates of the true effect, while appropriately controlling for false positives, with sufficient power ^23^. Linear mixed effects analyses were carried out using the lme4 package in R^24^ and robust regression was conducted using the robustbase package in R^25^. The meta-analysis was conducted using the metafor package for R^26^. Note that the meta-analysis did not take sex or age into account, but relied on the fact that the samples were approximately matched on these demographic variables in each individual study. The code used for the analyses is available here: https://github.com/ltuominen/SCRSCZ

## Results

### Group differences in Pavlovian fear conditioning

In the pooled sample, the difference between the SCRs to the CS+ and the CS-stimuli was significantly smaller in the schizophrenia, compared to the control, group (b = -0.134, t_143_ = -2.57, p = 0.011), (**Figure 1**; also see Figure 2 and Table 3 for data of the individual studies). The meta-analysis confirmed the findings of the linear mixed effects model (Cohen’s d = 0.5299, standard error = 0.1666, z = 3.1807, p = 0.0015, 95% CI = 0.2034 -0.8565). There was no evidence for heterogeneity across studies (I2 = 0%).

**Figure 1.**
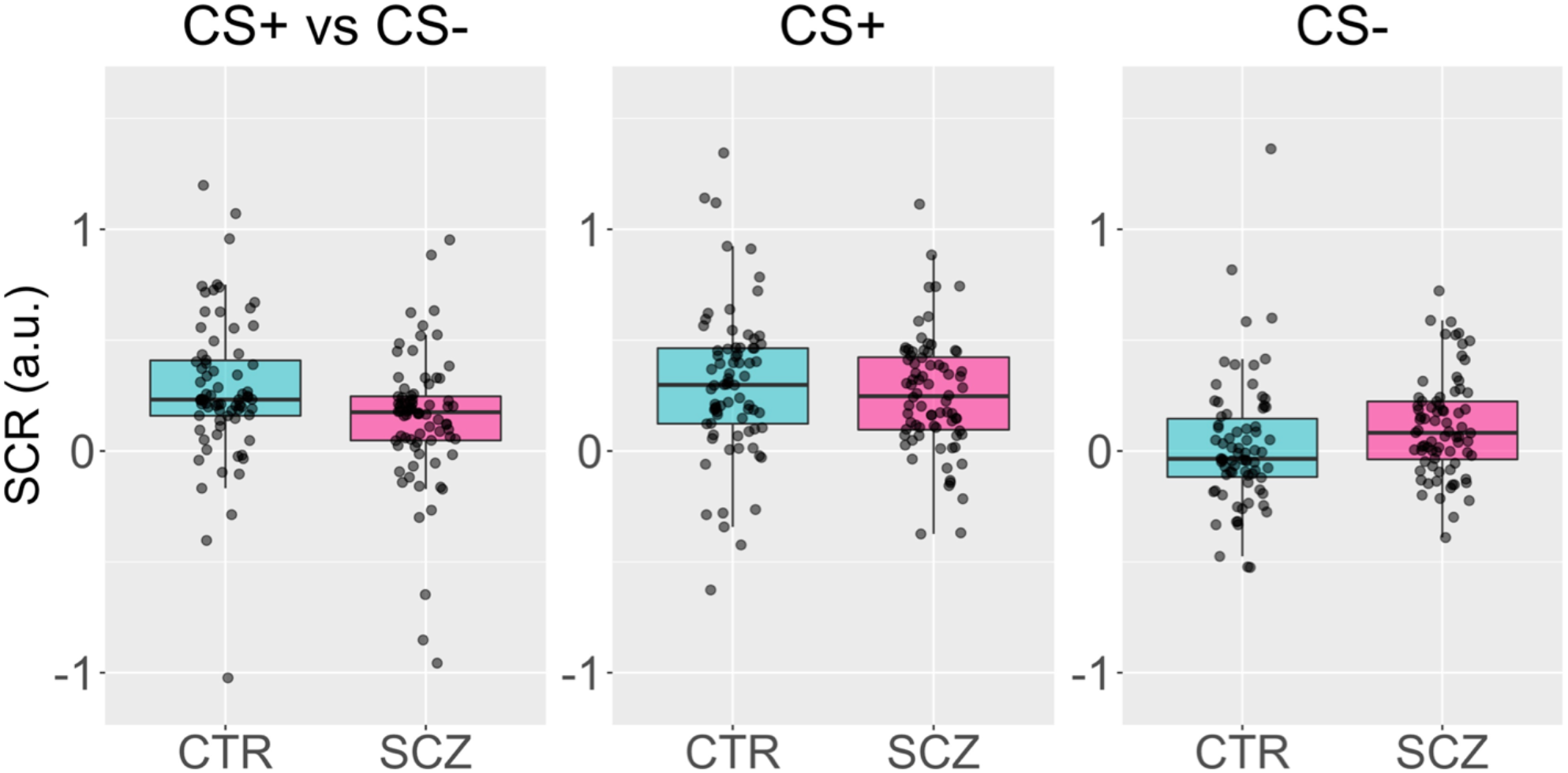
Acquisition of differential conditioned fear responses in 76 individuals with schizophrenia and 74 control subjects. The left panel shows that the difference between skin conductance responses (SCR) to the CS+ and CS- is lower in the schizophrenia (SCZ) than in the control (CTR) group (b = -0.134, t_143_ = -2.57, p = 0.011). The middle panel shows that SCR to the CS+ is on average numerically lower in SCZ than in CTR but this difference was not significant (b = -0.047, t_143_ = -0.966, p = 0.336). Finally, the right panel shows that the SCR to the CS- is significantly higher in SCZ compared to CTR (b = 0.087, t_143_ = 1.99, p = 0.048). For illustration, the individual datapoints have been adjusted for differences in age, sex and study.

**Figure 2.**
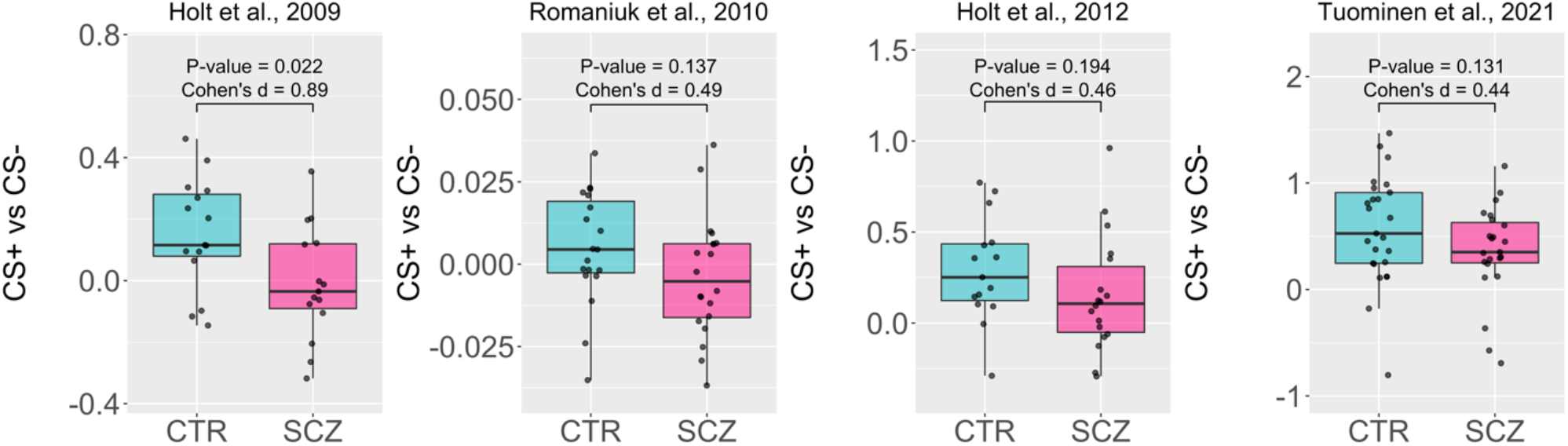
A pattern of diminished differential fear conditioning in schizophrenia is observed across studies. This figure shows that the mean difference between skin conductance responses to the CS+ and CS-stimuli is numerically smaller in the schizophrenia compared to the control group in each of the four studies included in the full analysis (but statistically significant only in Holt et al, 2009). Data presented in these graphs are not corrected for sex, age or study effects. CTR = control group, SCZ = schizophrenia group. As a preprocessing step, the Romaniuk et al., 2010 data were normalized to the maximum response and the scale is thus a magnitude smaller than in the other three studies.

**Table 3.** CS+ vs CS- contrast averages, standard deviations and group sizes in each study.

|  | Individuals with Schizophrenia |  |  | Control subjects |  |  |  |  |
| --- | --- | --- | --- | --- | --- | --- | --- | --- |
| Study | Contrast<br>Mean | Contrast<br>Std | N | Contrast<br>Mean | Contrast<br>Std | N | Effect size | Effect size<br>variance |
| HOLT<br>et al 2009 | -0.0095 | 0.184 | 15 | 0.1519 | 0.181 | 15 | 0.862 | 0.146 |
| ROMANIUK<br>et al 2010 | 0.1525 | 0.318 | 18 | 0.2927 | 0.287 | 15 | 0.449 | 0.125 |
| HOLT<br>et al 2012 | -0.0038 | 0.018 | 20 | 0.0048 | 0.017 | 19 | 0.476 | 0.106 |
| TUOMINEN<br>et al 2021 | 0.3558 | 0.441 | 23 | 0.5666 | 0.509 | 25 | 0.434 | 0.085 |

*Std* = standard deviation, *N* = number of participants

### Group differences in responses to the CS+ and CS-

The responses to the CS+ were on average slightly lower in the schizophrenia participants compared to the controls, but this difference was not statistically significant (b = -0.047, t_143_ = -0.966, p = 0.336), whereas the response to the CS- was significantly higher in the schizophrenia participants than in the controls (b = 0.087, t_143_ = 1.99, p = 0.048). These point estimates indicate that the increased response to the CS- (0.087) in the schizophrenia group accounted for 65% of the overall (0.134) decrease in the CS+ vs CS-contrast relative to the controls, while the non-significant decrease in response to the CS+ accounted for the remainder of the between-group difference.

### Correlations with clinical variables in the schizophrenia group

The PDI total score was positively correlated with the response to the CS- (b=0.333, t_50_=2.860, p=0.0062) (**Figure 3**). However levels of negative symptoms were not correlated with lower response to the CS+, nor were any other PANSS subscale or current antipsychotic dose correlated with any of the fear conditioning measures (**Table 4**).

**Figure 3.**
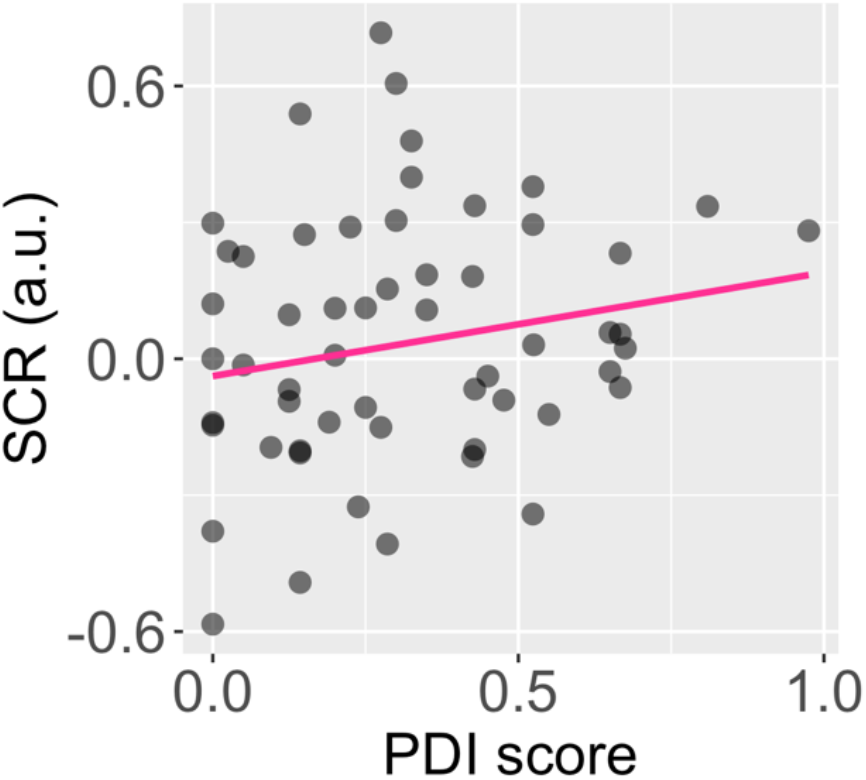
Delusion severity in schizophrenia is associated with a higher SCR response to the CS-. This plot shows the association between skin conductance responses (SCRs) to the CS-stimulus and the Peters Delusion Inventory total score (PDI total score), adjusted for the number of items, in 56 participants with schizophrenia. The regression line (b=0.333, t_50_=2.860, p=0.0062) has been adjusted for age, sex and study.

**Table 4.**
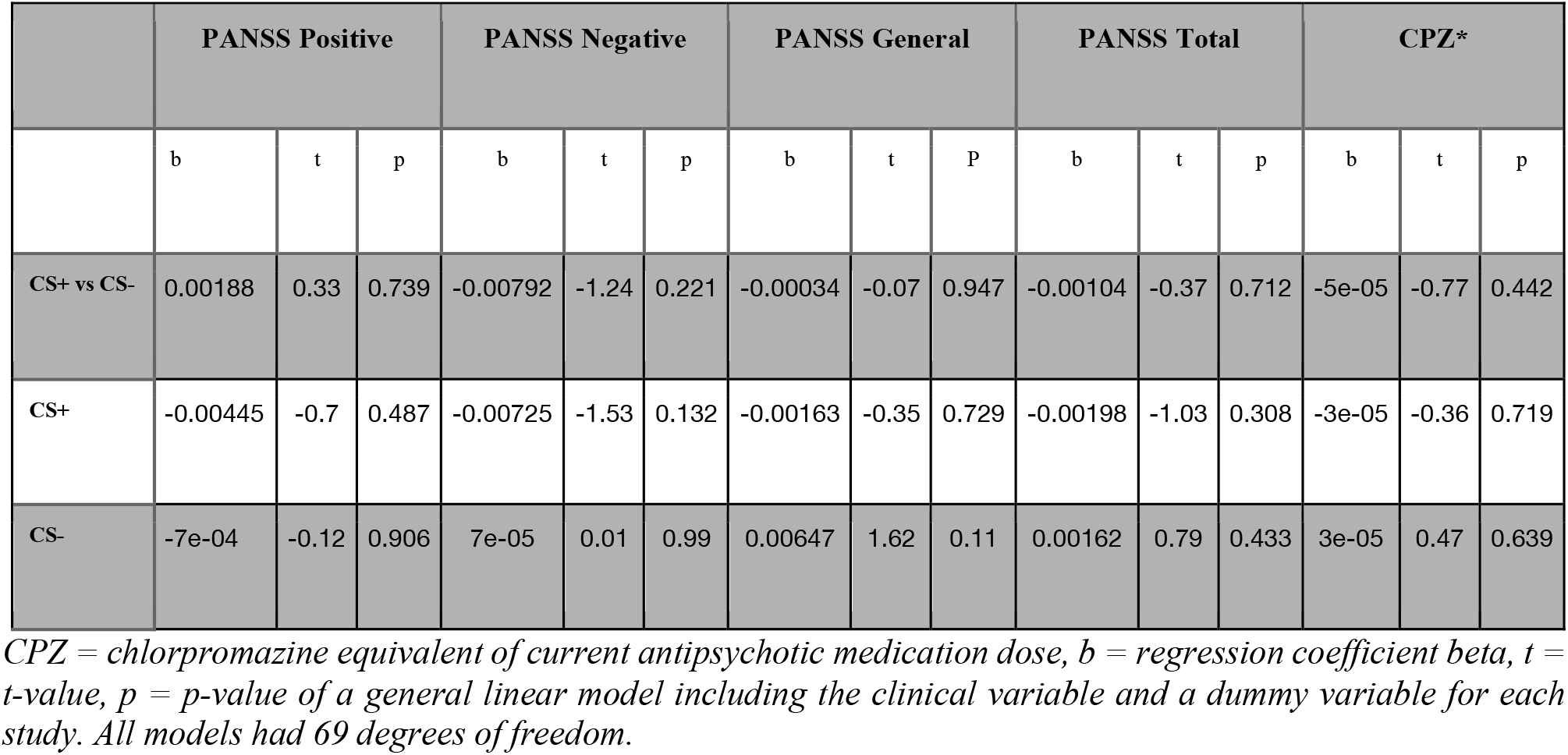
Results of the exploratory analyses testing for associations between clinical variables and skin conductance measures of fear conditioning in the schizophrenia group (n=76)

## Discussion

### Summary of findings

Here we show that acquisition of differential conditioned fear responses is impaired in schizophrenia using a pooled sample derived from four prior independent studies. The estimated effect size of schizophrenia diagnosis on fear acquisition in this sample (Cohen’s d = 0.53) was moderate and comparable to the effect of schizophrenia on hippocampal (Cohen’s d = 0.46)^27^ or lateral ventricle volumes (Cohen’s d = 0.37)^27^. Importantly, despite differences in the designs and paradigms of each study, there was no evidence for heterogeneity of the effect. Our analyses also revealed that the smaller difference between the CS+ and CS- was primarily due to higher responses to the CS-stimuli in the schizophrenia group, and that this response to the CS- was proportional to the severity of delusional ideation.

### Implicated neural circuitry

Prior studies using fMRI have tested this model, and some have reported increased BOLD responses to the CS- in the ventral striatum^6^, the midbrain^8^ and in the ventromedial prefrontal cortex^9,28^ in schizophrenia or clinical high-risk participants, in comparison to control subjects. In addition, an impaired ability to differentiate CS+ events from CS-events in schizophrenia is consistent with studies showing that schizophrenia is associated with diminished volumes of the amygdala ^27^, particularly since the lateral amygdala, where the association between the CS+ and US is learned, may be specifically affected^29,30^. Also, since the ventromedial prefrontal cortex exerts top-down control over the amygdala during emotional states^31^, disruption of the connections between the ventromedial prefrontal cortex and the amygdala in schizophrenia could lead to diminished inhibition of fear and greater responses to the CS-^32–34^.

### Fear conditioning in schizophrenia versus anxiety disorders

Individuals with anxiety disorders also tend to have slightly higher skin conductance responses to CS-stimuli during the fear acquisition phase^35^. Since responses to the CS+ are also higher in anxiety disorders, this finding has been thought to reflect overgeneralization of fear to the CS- events^35^. Curiously, we have shown that conditioned fear responses are undergeneralized in schizophrenia^12^. Overgeneralization is therefore unlikely to explain the elevated responses to the CS-in schizophrenia. In future work, identifying symptom-specific patterns in associative learning, specifically during fear conditioning, may facilitate the development of mechanism-based, targeted, early treatments of these specific symptom clusters.

### The potential role of dopamine in abnormalities in Pavlovian fear conditioning in schizophrenia

The aberrant salience attribution theory^3^ suggests that an increase in spontaneous dopamine transients heightens the salience of innocuous, neutral cues encountered in the environment, followed by a misinterpretation (and in some instances, a delusional explanation) of the experience. The CS-stimulus presented during Pavlovian fear conditioning paradigms may represent such a neutral cue in an experimental context ^6,8^. Accordingly, patients with schizophrenia who tend to have greater responses to the neutral, innocuous stimulus (the CS-) may be more likely to develop delusional ideas to account for this inappropriate salience signal. Under normal circumstances, dopamine is released in response to cues that predict either reward or punishment, which then guide associative learning^36^. Animal models and studies in humans have shown that the associative learning that occurs during Pavlovian fear conditioning is partially dopamine dependent. For instance, genetic deletion of dopamine D1 receptors in the striatum impairs this type of learning in mice^37^. In humans, a moderate dose of amphetamine increases the magnitude of fear conditioning responses^38^, presumably by increasing the level of adaptive dopamine release in response to predictive cues^39^. However, at higher doses, amphetamine increases the spontaneous dopamine transients that are not associated with predictions, while blunting the adaptive dopamine responses^39^. Based on these data, we speculate that the elevated responses to the CS-observed in schizophrenia in the current study is attributable to such an increase in spontaneous dopamine release, which may persist to an extent even in individuals treated with antipsychotic medication. Moreover, our finding that higher skin conductance responses to the CS- is associated with higher scores on the Peters Delusion Inventory is consistent with this model of psychosis.

While the response to the CS+ was on average numerically lower in the schizophrenia group in the current study, there were no statistically significant between-group differences in responses to the CS+. Yet, the point estimate of increased response to the CS- did not entirely explain the group difference in the acquisition of conditioned fear; some residual deficit may be attributable to a decreased response to the CS+ in the schizophrenia group. The response to the CS+ is thought to reflect cue-induced adaptive dopamine release^7^, similar to prediction error learning related dopamine release^45^. Studies using reinforcement learning paradigms have shown that while deficits in learning from rewards is associated with negative symptoms, learning from aversive outcomes may be spared in schizophrenia^41^. Those prior results are consistent with our current findings of relatively intact responses to the CS+ (a negative reinforcer). Finally, changes in other neurotransmitters, such as glutamate^42,43^, acetylcholine^44,45^ and serotonin^46^, which play a role in fear conditioning, may also contribute to the impairments observed in schizophrenia in associative learning.

### Limitations

Several limitations of this pooled analysis include those of the original studies, such as the fact that the majority of the individuals with schizophrenia enrolled were receiving treatment with antipsychotic medication. Treatment with antipsychotic medications, which are typically D2 dopamine receptor antagonists, may affect fear conditioning^38^. However, antipsychotic dose was not correlated with any of the skin conductance measures in the schizophrenia sample. Nevertheless, adequately powered future studies should investigate whether unmedicated individuals with schizophrenia display similar deficits in fear acquisition. Another limitation of this pooled analysis is that one recent Pavlovian fear conditioning study of 10 individuals with schizophrenia and 11 healthy controls was not included because the original data were no longer available^6^. However, the findings of this study were in line with the results reported here, i.e., the schizophrenia group showed a significantly smaller difference between SCRs to the CS+ and CS-, due to both lower response to the CS+ and higher response to the CS-.

### Conclusions

In a pooled sample derived from four independent studies, we found that schizophrenia is associated with impaired acquisition of differential fear conditioning, due to an increased response to the neutral stimulus. Moreover, this elevated response to the neutral stimulus was proportional to the severity of delusional ideation. Fear conditioning is a simple experimental paradigm which could be used in future work to establish the mechanisms associated with psychotic symptoms and how they manifest during the early course of the illness.

## Data Availability

Original anonymized data is available upon reasonable request.

https://github.com/ltuominen/SCRSCZ

